# Greater medial temporal lobe atrophy is associated with less severe cardiac sympathetic denervation in dementia with Lewy bodies

**DOI:** 10.64898/2026.11.13.26367891

**Authors:** Makoto Takahashi, Narumi Kitsuka, Hiroyuki Maruoka, Keisuke Abe, Satoshi Orimo, Akira Inaba

## Abstract

**Background:** Cardiac sympathetic denervation varies in dementia with Lewy bodies (DLB). Our previous neuropathological study demonstrated an association between greater Alzheimer’s disease (AD) pathology and milder cardiac sympathetic denervation in DLB. We investigated whether medial temporal lobe atrophy (MTA), a potential marker of concomitant AD pathology, correlates with cardiac sympathetic denervation assessed by ^123^I-metaiodobenzylguanidine (MIBG) cardiac scintigraphy in probable DLB.

**Methods:** We retrospectively analyzed 162 patients with probable DLB. MTA was assessed by VSRAD Z-scores, and cardiac MIBG uptake by early and delayed heart-to-mediastinum (H/M) ratios. Associations were examined using Spearman and partial Spearman correlations adjusted for age at diagnosis, sex, and disease duration. Sensitivity analyses excluded patients with MIBG-dependent classification, diabetes mellitus or cardiac disease.

**Results:** VSRAD Z-scores correlated positively with early (ρ = 0.173, P = 0.028) and delayed H/M ratios (ρ = 0.225, P = 0.004). After adjustment, the association was no longer significant for the early H/M ratio (ρ = 0.133, P = 0.094), whereas it remained significant for the delayed H/M ratio (ρ = 0.196, P = 0.013). The adjusted delayed H/M association persisted after excluding 18 patients with MIBG-dependent classification (ρ = 0.211, P = 0.012) and 40 with diabetes or cardiac disease (ρ = 0.231, P = 0.011).

**Conclusions:** Greater MTA was associated with relatively preserved cardiac MIBG uptake in probable DLB, particularly on delayed imaging. Consistent with our neuropathological observations, these findings suggest that concomitant AD pathology may contribute to variability in cardiac sympathetic denervation.

## Introduction

Dementia with Lewy bodies (DLB) is a neurodegenerative dementia characterized by Lewy body pathology composed predominantly of aggregated α-synuclein. Relative preservation of medial temporal lobe structures is a supportive neuroimaging biomarker of DLB [1]. However, Alzheimer’s disease (AD) pathology frequently coexists in patients with DLB, and its presence is associated with greater atrophy of the hippocampus and other medial temporal structures [2,3]. Thus, the degree of medial temporal lobe atrophy (MTA) may partly reflect the pathological heterogeneity of DLB.

Cardiac sympathetic denervation is another characteristic feature of Lewy body diseases [1,4]. Reduced cardiac uptake of ^123^I-metaiodobenzylguanidine (MIBG) reflects degeneration of postganglionic cardiac sympathetic nerves and is an established indicative biomarker for the clinical diagnosis of DLB [5]. However, the extent of cardiac sympathetic denervation varies among patients with DLB. In our previous neuropathological study of autopsy-confirmed DLB, patients with substantial concomitant AD pathology had relatively preserved cardiac sympathetic nerve axons, suggesting that the degree of cardiac sympathetic denervation may differ according to the underlying pathological profile of DLB [6].

These findings raise the possibility that greater MTA is associated with less severe cardiac sympathetic denervation in DLB. However, the relationship between MTA and cardiac MIBG uptake in clinically diagnosed DLB has not been well established. Therefore, we investigated the association between MTA assessed using the Voxel-based Specific Regional Analysis System for Alzheimer’s Disease (VSRAD) and cardiac MIBG uptake in patients with probable DLB. We hypothesized that greater MTA would be associated with relative preservation of cardiac MIBG uptake.

## Materials and Methods

### Ethics Statement

The study was approved by the Institutional Review Board of Kanto Central Hospital (approval number: R6-6). Because of the retrospective nature of the study, participants were provided with the opportunity to opt out of the study.

### Study Design and Participants

This retrospective observational study screened patients at the Department of Neurology, Kanto Central Hospital, between April 1, 2015, and March 31, 2024. Patients who underwent both brain magnetic resonance imaging (MRI) with VSRAD analysis and MIBG cardiac scintigraphy within one month of each other were screened.

Clinical information was retrospectively obtained from the medical records. The diagnosis of probable DLB was determined by applying the 2017 revised consensus criteria of the DLB Consortium to the available clinical and imaging data [1]. Core clinical features, including cognitive fluctuations, visual hallucinations, parkinsonism, and rapid eye movement sleep behavior disorder (RBD), were identified from documented clinical histories obtained from patients and/or caregivers. Patients who fulfilled the criteria for probable DLB were included in the analysis.

### ^123^I-MIBG cardiac scintigraphy

After intravenous administration of 128 MBq of ¹²³I-MIBG, planar images were obtained at 30 min (early phase) and 3 h (delayed phase). Early and delayed heart-to-mediastinum (H/M) ratios were calculated using semiautomated software. Cardiac MIBG scintigraphy was not performed in patients receiving medications known to interfere with MIBG uptake.

### Assessment of medial temporal lobe atrophy

MTA was quantitatively assessed using the VSRAD (Eisai Co., Ltd., Tokyo, Japan) as part of routine clinical brain MRI evaluation. MRI examinations were performed using a 1.5-T Siemens MAGNETOM Symphony system or a 3-T Siemens MAGNETOM Verio system. The VSRAD Z-score, which represents the severity of gray matter atrophy in the medial temporal region relative to a normal reference database, was used as an index of MTA, with higher Z-scores indicating greater atrophy [7].

### Statistical Analysis

All statistical analyses were performed using EZR [8]. Continuous variables were expressed as means ± standard deviations or medians with interquartile ranges (IQRs), as appropriate, and categorical variables as numbers and percentages. Associations between VSRAD Z-scores and cardiac MIBG H/M ratios were assessed using Spearman’s rank correlation coefficients. Age at diagnosis, sex, and disease duration were selected as covariates based on previous reports suggesting potential associations with cardiac MIBG uptake [6,9,10]. Partial Spearman’s rank correlation analyses were performed to adjust for these covariates, with two-sided P values obtained using a t-distribution approximation. To address potential circularity related to the use of reduced cardiac MIBG uptake in the diagnosis of probable DLB, a sensitivity analysis was performed after excluding patients whose diagnosis of probable DLB depended on reduced cardiac MIBG uptake. A separate sensitivity analysis was performed after excluding patients with diabetes mellitus or cardiac disease (myocardial infarction, heart failure, or cardiomyopathy), as these conditions may independently affect cardiac MIBG uptake [11,12]. Spearman correlation and partial Spearman correlation analyses were repeated in each restricted cohort. All statistical tests were two-sided, and P values <0.05 were considered statistically significant. There were no missing data for any variables included in the correlation analyses.

## Results

### Patient characteristics

A total of 162 consecutive patients with probable DLB were included in the analysis (Table 1). The mean age at diagnosis was 82.8 ± 5.7 years, and 71 patients (43.8%) were male. The median disease duration was 1.57 years (interquartile range [IQR], 0.67–3.17). The median Mini-Mental State Examination (MMSE) score was 23 (IQR, 19–25); MMSE data were available for 135 patients. Cognitive fluctuations were present in 59 patients (36.4%), visual hallucinations in 123 (75.9%), RBD in 82 (50.6%), and parkinsonism in 118 (72.8%). Diabetes mellitus was present in 29 patients (17.9%), and a history of cardiac disease was identified in 17 patients (10.5%); six patients (3.7%) had both conditions. The median early and delayed cardiac MIBG H/M ratios were 1.72 (IQR, 1.52–2.09) and 1.44 (IQR, 1.25–1.76), respectively. The median VSRAD Z-score was 1.65 (IQR, 1.17–2.40). Associations of age at diagnosis, sex, and disease duration with VSRAD Z-scores and cardiac MIBG H/M ratios are summarized in Table S1.

**Table 1.** Clinical characteristics of the study population. Data are presented as mean ± SD, median [IQR], or n (%), as appropriate. MMSE data were available for 135 patients. Cardiac disease was defined as myocardial infarction, heart failure, or cardiomyopathy. Abbreviations: MMSE, Mini-Mental State Examination; RBD, rapid eye movement sleep behavior disorder; MIBG, ^123^I-metaiodobenzylguanidine; H/M, heart-to-mediastinum; VSRAD, voxel-based specific regional analysis system for Alzheimer’s disease.

| Characteristics | Overall (n=162) |
|---|---|
| Male sex, n (%) | 71 (43.8) |
| Age at onset, years | 80.5 ± 6.1 |
| Age at diagnosis, years | 82.8 ± 5.7 |
| Disease duration, years | 1.57 [0.67–3.17] |
| MMSE score | 23 [19–25] |
| Cognitive fluctuations, n (%) | 59 (36.4) |
| Visual hallucinations, n (%) | 123 (75.9) |
| RBD, n (%) | 82 (50.6) |
| Parkinsonism, n (%) | 118 (72.8) |
| Diabetes mellitus, n (%) | 29 (17.9) |
| Cardiac disease, n (%) | 17 (10.5) |
| MIBG early H/M ratio | 1.72 [1.52–2.09] |
| MIBG delayed H/M ratio | 1.44 [1.25–1.76] |
| VSRAD Z-score | 1.65 [1.17–2.40] |

### Association between MTA and cardiac MIBG uptake

The VSRAD Z-score was positively correlated with both the early and delayed cardiac MIBG H/M ratios. Spearman’s rank correlation coefficients were ρ = 0.173 (P = 0.028) for the early H/M ratio and ρ = 0.225 (P = 0.004) for the delayed H/M ratio (Figure 1).

**Figure 1.**
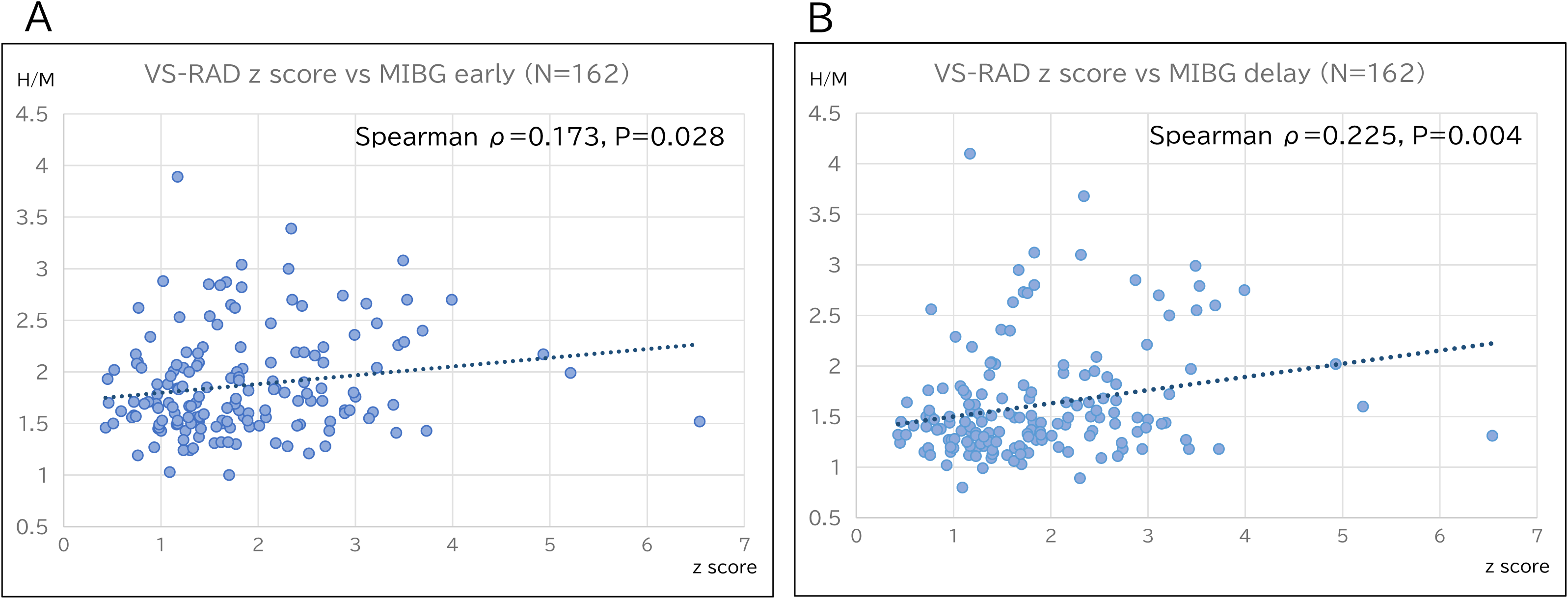
Associations between medial temporal lobe atrophy and cardiac MIBG uptake in patients with dementia with Lewy bodies. Scatterplots showing the associations between VSRAD Z-scores and (A) early and (B) delayed cardiac MIBG heart-to-mediastinum (H/M) ratios in 162 patients with probable dementia with Lewy bodies. Spearman’s rank correlation coefficients were ρ = 0.173 (P = 0.028) for the early H/M ratio and ρ = 0.225 (P = 0.004) for the delayed H/M ratio.

After adjustment for age at diagnosis, sex, and disease duration using partial Spearman correlation analysis, the association was no longer significant for the early H/M ratio (ρ = 0.133, P = 0.094), whereas it remained significant for the delayed H/M ratio (ρ = 0.196, P = 0.013). To address potential diagnostic circularity, we performed a sensitivity analysis excluding 18 patients whose classification as probable DLB depended on reduced cardiac MIBG uptake. Among the remaining 144 patients, the association between the VSRAD Z-score and delayed H/M ratio remained significant (Spearman’s ρ = 0.240, P = 0.004). The association also remained significant after adjustment for age at diagnosis, sex, and disease duration (partial Spearman’s ρ = 0.211, P = 0.012). In a separate sensitivity analysis excluding 40 patients with diabetes mellitus or cardiac disease, 122 patients remained. The VSRAD Z-score was positively correlated with both the early (Spearman’s ρ = 0.227, P = 0.012) and delayed H/M ratios (ρ = 0.263, P = 0.003). After adjustment for age at diagnosis, sex, and disease duration, the associations remained significant for both the early (partial Spearman’s ρ = 0.198, P = 0.031) and delayed H/M ratios (ρ = 0.231, P = 0.011) (Table 2).

**Table 2.**
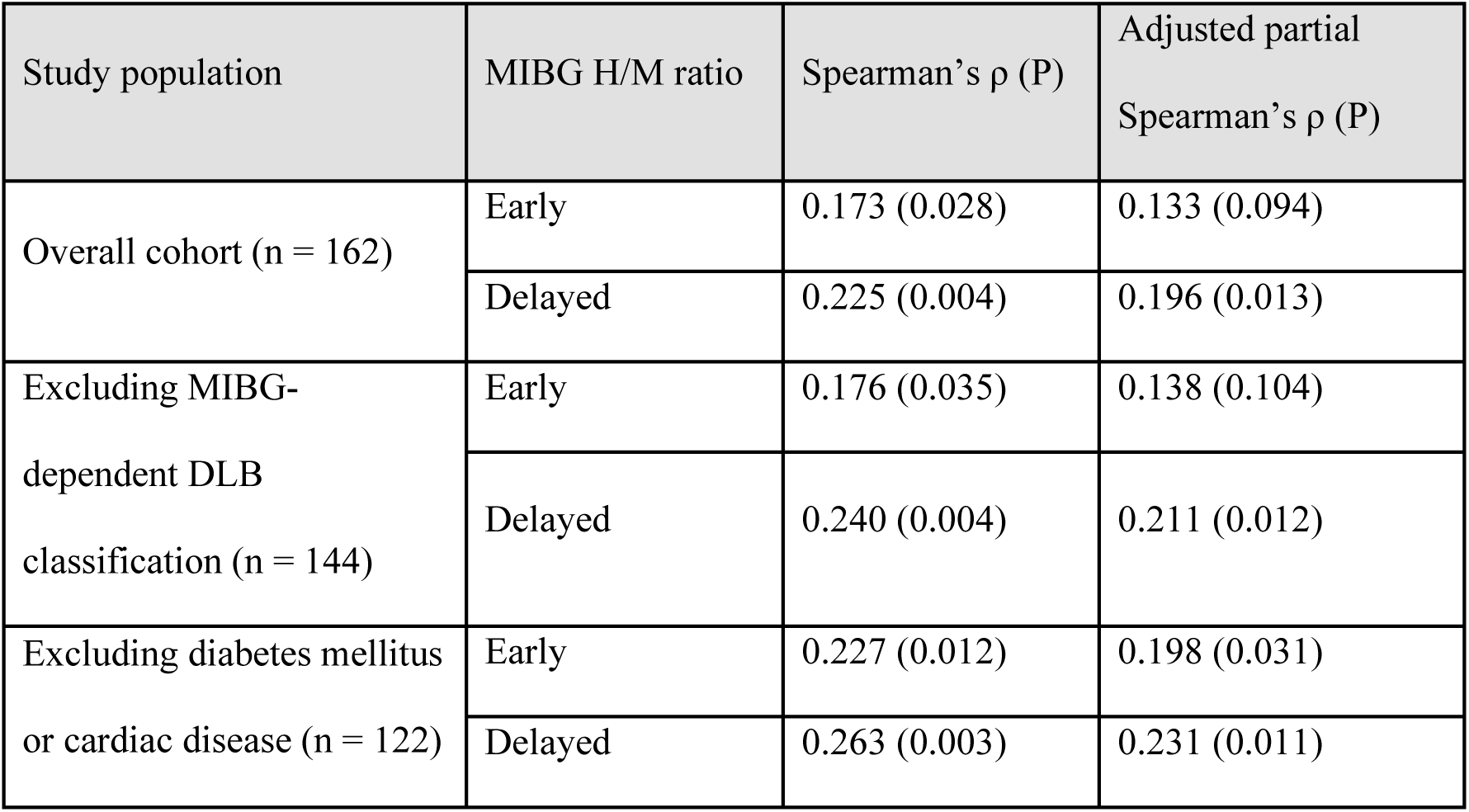
Associations between VSRAD Z-scores and cardiac MIBG H/M ratios in the overall cohort and sensitivity analyses. Values are Spearman’s rank correlation coefficients or partial Spearman’s rank correlation coefficients, with P values in parentheses. Partial correlations were adjusted for age at diagnosis, sex, and disease duration. The sensitivity analyses were performed separately. Abbreviations: DLB, dementia with Lewy bodies; H/M, heart-to-mediastinum; MIBG, ¹²³I-metaiodobenzylguanidine; VSRAD, Voxel-based Specific Regional Analysis System for Alzheimer’s Disease.

## Discussion

In the present study, greater MTA, as assessed by the VSRAD Z-score, was associated with relatively preserved cardiac MIBG uptake in patients with clinically diagnosed probable DLB. The association remained significant for the delayed H/M ratio after adjustment for age, sex, and disease duration. Furthermore, this association persisted in two separate sensitivity analyses: one excluding patients whose classification as probable DLB depended on reduced cardiac MIBG uptake, and another excluding those with diabetes mellitus or cardiac disease. These findings suggest that greater MTA is associated with less severe cardiac sympathetic denervation in DLB.

The present findings are consistent with our previous neuropathological observations. In our previous study of autopsy-confirmed DLB, cardiac sympathetic nerve fibers were relatively preserved in patients with substantial concomitant AD pathology, suggesting an inverse relationship between the severity of AD pathology and cardiac sympathetic denervation [6]. Importantly, our earlier clinicopathological study of autopsy-confirmed LBD demonstrated that cardiac MIBG H/M ratios were quantitatively correlated with the extent of residual cardiac sympathetic axons, supporting cardiac MIBG uptake as an in vivo marker of cardiac sympathetic denervation [5]. Thus, the present association between greater MTA and relatively preserved cardiac MIBG uptake may represent a clinical correlate of our previous neuropathological findings.

Relative preservation of medial temporal lobe structures is a characteristic imaging feature of DLB compared with AD [1]. However, DLB is pathologically heterogeneous, and concomitant AD pathology is common. Previous neuropathologically verified studies have demonstrated associations of medial temporal lobe atrophy and its progression with AD-related pathology, particularly neurofibrillary tangle pathology [2,3]. Although the VSRAD Z-score is not a direct measure of AD pathology, greater MTA in the present study may partly reflect concomitant AD-related neurodegeneration. Nevertheless, the observed correlations were weak, with substantial variability in cardiac MIBG uptake across the range of VSRAD Z-scores (Figure 1). These findings suggest that MTA alone provides limited information about the severity of cardiac sympathetic denervation in individual patients with DLB.

One possible explanation for the observed association is that concomitant AD pathology may influence the clinical expression of DLB. Patients with greater AD-related neurodegeneration may reach the clinical threshold for dementia at an earlier stage of Lewy body-related neurodegeneration, potentially before extensive cardiac sympathetic denervation has occurred. Conversely, patients with less concomitant AD pathology may require more advanced Lewy body-related neurodegeneration before dementia becomes clinically apparent, resulting in more pronounced cardiac sympathetic denervation at the time of diagnosis. This hypothesis provides a possible explanation for the inverse relationship between concomitant AD pathology and cardiac sympathetic denervation observed in our previous neuropathological study [6].

However, the present cross-sectional findings cannot establish the temporal sequence of central neurodegeneration and peripheral sympathetic denervation. From a clinical perspective, these findings suggest that relatively preserved cardiac MIBG uptake in patients with suspected DLB and marked MTA should be interpreted in the context of possible concomitant AD-related neurodegeneration, rather than being regarded as evidence against DLB in isolation.

The association with MTA remained significant after adjustment for age, sex, and disease duration for the delayed H/M ratio, but not for the early H/M ratio in the full cohort. This finding is consistent with our previous clinicopathological study of autopsy-confirmed LBD, in which the delayed H/M ratio showed slightly stronger correlations with residual cardiac sympathetic axons than the early H/M ratio [5]. These observations suggest that delayed MIBG uptake may be particularly informative for assessing cardiac sympathetic denervation in LBD. However, the differences between early and delayed imaging were small, and the present study did not directly compare their associations with cardiac sympathetic pathology.

Several limitations should be acknowledged. First, this was a retrospective, single-center study, and the diagnosis of DLB was based on clinical information rather than neuropathological confirmation. However, the association between MTA and delayed MIBG uptake remained significant after excluding patients whose classification as probable DLB depended on reduced MIBG uptake, reducing the possibility that the observed association resulted from circularity in case ascertainment. In addition, only patients who underwent both brain MRI with VSRAD analysis and cardiac MIBG scintigraphy were included, potentially introducing selection bias. Therefore, the generalizability of our findings to the broader DLB population may be limited. Second, core clinical features of DLB, including cognitive fluctuations, visual hallucinations, and RBD, were identified retrospectively from documented clinical histories rather than through standardized assessments. In particular, polysomnography and validated RBD questionnaires were not systematically available. Third, biomarkers of AD pathology, such as amyloid or tau biomarkers, and neuropathological confirmation were not available in this study. Although greater MTA may reflect concomitant AD-related neurodegeneration, MTA is not specific to AD pathology and may also be influenced by aging and other neurodegenerative processes. Therefore, the contribution of AD pathology to the observed association remains speculative.

In conclusion, greater MTA was associated with relatively preserved cardiac MIBG uptake, particularly on delayed imaging, in patients with probable DLB. This finding is consistent with our previous neuropathological observations suggesting relative preservation of cardiac sympathetic innervation in DLB with substantial concomitant AD pathology.

## Acknowledgments

The authors would like to thank Mr. Jun Amano for his technical support in MRI acquisition and VSRAD analysis.

## Author Contributions

Conceptualization: MT

Formal analysis: MT

Investigation: MT, NK, HM, KA, AI

Data curation: MT, AI

Writing-original draft: MT

Writing-review & editing: MT, SO, AI

Supervision: SO, AI

## Data Availability

The data that support the findings of this study are available from the corresponding author upon reasonable request.

## Declaration of generative AI and AI-assisted technologies in the manuscript preparation process

During the preparation of this work, the authors used ChatGPT (OpenAI) to assist with language editing, manuscript organization, and the development and checking of statistical analysis code. After using this tool, the authors reviewed and edited the content as needed and take full responsibility for the content of the published article.

