## Supplemental Table 1 for "Greater medial temporal lobe atrophy is associated with less severe cardiac sympathetic denervation in dementia with Lewy bodies"

Table S1. Associations of age, disease duration, and sex with VSRAD Z-scores and cardiac MIBG H/M ratios in the full cohort (n = 162).

A. Correlations with age and disease duration

| Variable | Age at diagnosis, ρ (P) | Disease duration, ρ (P) |
|---|---|---|
| VSRAD Z-score | 0.167 (0.034) | 0.212 (0.007) |
| Early H/M ratio | 0.037 (0.638) | 0.193 (0.014) |
| Delayed H/M ratio | 0.004 (0.957) | 0.188 (0.017) |

B. Comparisons by sex

| Variable | Female (n = 91) | Male (n = 71) | P value |
|---|---|---|---|
| VSRAD Z-score | 1.67 [1.16–2.40] | 1.58 [1.19–2.38] | 0.972 |
| Early H/M ratio | 1.83 [1.53–2.17] | 1.66 [1.51–2.00] | 0.085 |
| Delayed H/M ratio | 1.49 [1.29–1.85] | 1.39 [1.22–1.65] | 0.094 |

Values in panel A are Spearman's rank correlation coefficients (ρ), with P values in parentheses. Values in panel B are medians [interquartile ranges], with comparisons performed using the Mann–Whitney U test. All P values are two-sided and unadjusted for multiple comparisons.

Abbreviations: VSRAD, Voxel-based Specific Regional Analysis System for Alzheimer's Disease; MIBG, $^{123}$I-metaiodobenzylguanidine; H/M, heart-to-mediastinum.
